# Predicting the impact of COVID-19 non-pharmaceutical intervention on short- and medium-term dynamics of enterovirus D68 in the US

**DOI:** 10.1101/2023.07.14.23292680

**Authors:** Sang Woo Park, Kevin Messacar, Daniel C. Douek, Alicen B. Spaulding, C. Jessica E. Metcalf, Bryan T. Grenfell

## Abstract

Recent outbreaks of enterovirus D68 (EV-D68) infections, and their causal linkage with acute flaccid myelitis (AFM), continue to pose a serious public health concern. During 2020 and 2021, the dynamics of EV-D68 and other pathogens have been significantly perturbed by non-pharmaceutical interventions against COVID-19; this perturbation presents a powerful natural experiment for exploring the dynamics of these endemic infections. In this study, we analyzed publicly available data on EV-D68 infections, originally collected through the New Vaccine Surveillance Network, to predict their short- and long-term dynamics following the COVID-19 interventions. Although there are large uncertainties in our predictions, the likelihood of a large outbreak in 2023 appears to be low. Comprehensive surveillance data are needed to narrow uncertainties in future dynamics of EV-D68. The limited incidence of AFM cases in 2022, despite large EV-D68 outbreaks, poses further questions for the timing of the next AFM outbreaks.

## 1 Introduction

Enterovirus D68 (EV-D68) is a childhood infection typically associated with respiratory symptoms [1]. Even though it was first discovered in 1968 [2], cases were reported sporadically until 2014 when a large outbreak occurred in the United States (US) [3] and many other countries. Around this period, a novel neurological disease exhibiting polio-like symptoms—now referred to as acute flaccid myelitis (AFM)—also emerged [4, 5]. Accumulating evidence supporting a causal link between EV-D68 infections and AFM has raised public health concerns across the world. EV-D68 alone also exerts a significant direct burden as a widespread childhood infection. Both these factors highlight the necessity to understand its past and future dynamics [6].

Between 2014–2018, EV-D68 outbreaks were reported every two years in the US (with local variations) and in many other countries [7]. Therefore, many anticipated an EV-D68 outbreak in 2020; in particular, our previous modeling analysis predicted that an outbreak would be likely across a wide range of parameter regimes [8]. However, as COVID-19 emerged in early 2020, non-pharmaceutical interventions (NPIs) were introduced, thereby reducing contact rates and preventing the transmission of endemic diseases across the world. We also predicted that even a modest reduction in contact rates would prevent an EV-D68 outbreak in 2020 [8]. Analyses of other respiratory pathogens, such as RSV and influenza, further predicted that the accumulation of the susceptible pool during the NPI period would result in a larger outbreak when NPIs were relaxed [9].

As predicted, EV-D68 circulations remained low internationally in 2020 and large outbreaks were reported in following years with some heterogeneity. In Europe, EV-D68 outbreaks were primarily reported in 2021 [10, 11]. In the US, EV-D68 outbreaks were reported in 2022 in many places [12] with some reportings in 2021 as well [13]. However, even as EV-D68 re-emerged, there was limited detection of AFM cases; this adds uncertainty to the etiology to the uncoupling of their dynamics and the predictions for future outbreaks.

In this study, we analyzed the spread of EV-D68 in the US between 2018 and 2022 using previously published data collected from the Centers for Disease Control and Prevention (CDC) New Vaccine Surveillance Network (NVSN) [12]. We used mathematical models to estimate underlying parameters and characterize the impact of non-pharmaceutical interventions on the future dynamic of EV-D68. We further performed a series of sensitivity analyses to capture the uncertainty in long-term predictions for future EV-D68 outbreaks in the US.

## 2 Methods

### 2.1 Data

We digitized publicly available syndromic surveillance and viral testing data between 2018 and 2022 from the figure of [12] using https://automeris.io/WebPlotDigitizer/. The original paper presents three time series data: percentages of emergency department (ED) visits for children and adolescents (*<* 18 years old) associated with acute respiratory illness (ARI), percentages of positives for rhinovirus (RV) and enterovirus (EV), and percentages of positives for EV-D68 among RV/EV positives. Emergency department visits were originally collected through the National Syndromic Surveillance Program. Rhinovirus and enterovirus testing data were originally collected through the National Respiratory and Enteric Virus Surveillance System. Finally, EV-D68 testing data were originally collected through the New Vaccine Surveillance Network (NVSN). In this study, we rely on published graphical summaries of three data sets. Due to the sparsity of EV-D68 data, we only digitized data points that could be distinguished easily from visual inspection.

To calculate an incidence proxy for EV-D68, we multiplied weekly percentages of ED visits associated with ARIs with weekly percentages of RV/EV positives and weekly percentages of EV-D68 positives. As discussed previously in many contexts [14, 15], positivity rates can give a biased perspective on viral circulation levels because they are also sensitive to circulations of other pathogens exhibiting similar symptoms. Instead, this incidence proxy would accurately capture the dynamics of infections up to a constant under four assumptions explained in [14]. To validate the robustness of this proxy, we also tried using the weekly proportions of influenza-like illness (ILI) visits, instead of ED visits associated with ARIs, to calculate the incidence proxy. ILI data were downloaded from the Outpatient Influenza-like Illness Surveillance Network (ILINet). All data are presented in Supplementary Figure S1. As a comparison, we considered the monthly EV-D68 positivity data collected through BioFire for the period 2014–2019. The data are publicly available from [8]. We did not have access to data for RV/EV positivity or ED visits during this period and so we were not able to calculate the incidence proxy. We excluded this data from model fitting for simplicity.

To capture the dynamics of COVID-19 NPIs, we also analyzed mobility data collected from Google. Since only the most recent data are publicly available through Google (https://www.google.com/covid19/mobility/), we downloaded the mobility data from Our World in Data instead (https://ourworldindata.org/covid-google-mobility-tends). We calculated the weekly mean mobility across four categories: retail & recreation, grocery & pharmacy, transit stations, and workplaces.

### 2.2 Model

We used the deterministic SIR model to capture the spread of EV-D68:

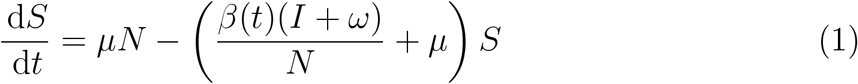

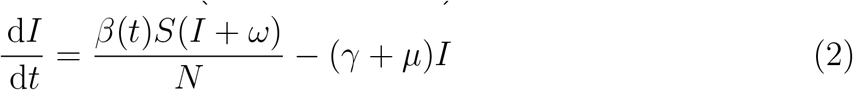

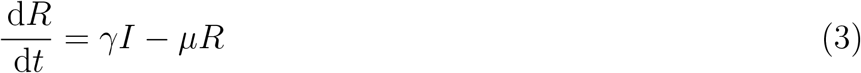

where state variables *S, I*, and *R* represent the number of susceptible, infected, and recovered individuals; *N* = *S* +*I* +*R* represent the total population size; *µ* represents the birth and death rate; *β* represents the transmission rate; *ω* represents the rate of external infections; and *γ* represents the recovery rate. Previous studies showed that an SIR model, assuming predominantly strong and durable transmission-blocking immunity from primary infection, can capture the dynamics of many enterovirus species, including EV-D68 [16, 8]. The incidence *i*(*t*) was calculated by taking the differences between the cumulative infections *i*(*t*) = *C*(*t*) − *C*(*t* − 1), where:

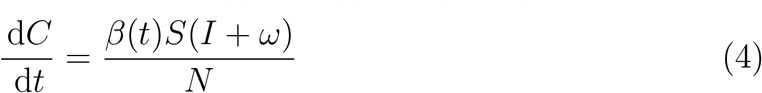

For simplicity, we assumed a fixed value of birth/death rate 1/*µ* = (80×52) weeks and recovery rate 1/*γ* = 1 week throughout this paper. We also assumed a population size of 1 million and scale the incidence appropriately for model fitting.

We further extended the SIR model to allow for changes in the contact rates during the pandemic period and seasonal variation in transmission:

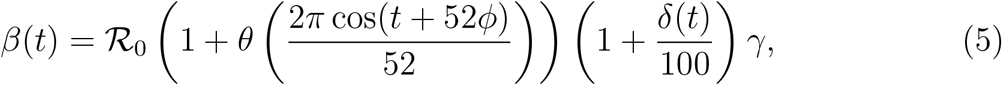

where ℛ_0_ represents the basic reproduction number; *θ* represents the amplitude of seasonal forcing; *ϕ* represents the offset parameter; and *δ*(*t*) represents the percent change in contact rates.

We considered two sets of assumptions for modeling the dynamics of EV-D68 and the impact of NPIs. For the first set of assumptions, we assumed that the EV-D68 was following an endemic biennial pattern. Based on previous parameter estimates, we assumed ℛ_0_ = 20, *θ* = 0.2, and *ω* = 2, which sits in a stable biennial regime. By varying the parameter *φ* between 0.3 and 0.5 for each simulation, we ran the model for 24 years prior to 2018 starting from the initial conditions *S*(0) = (1/ ℛ_0_)*N, I*(0) = 1, and *R*(0) = *N* −*S*(0) −*I*(0) such that the 25th year in the simulation corresponds to 2018. Then, we tried to find a parameter set that maximized the likelihood assuming normally distributed errors:

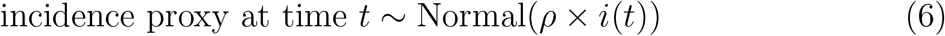

where *ρ* represents the scaling factor. For each simulation, we performed a linear regression between incidence proxy and the predicted incidence for 2018 without the intercept term, which allowed us to calculate the likelihood as well as the scaling factor (corresponding to the estimated coefficient).

Using the best matching seasonality term *ϕ*, we further tried to estimate the changes in contact rates *δ*(*t*) that best matches the data. We initially explored whether a constant reduction in contact rate for a fixed amount of time alone can explain the observed dynamics:

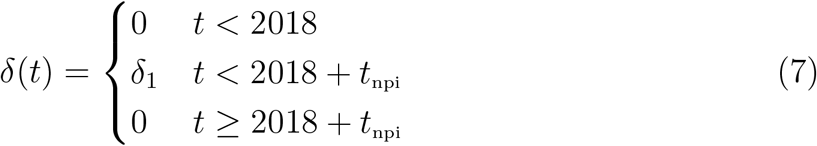

where *δ*_1_ represents the amount of reduction in contact rates, and *t*_npi_ represents the duration of non-pharmaceutical interventions. During this investigation, we found that this simple functional form was not able to capture the observed dynamics sufficiently, and instead overestimated the size of the epidemic in 2022 (Supplementary Figure S2). Therefore, we instead assumed that contact rates were initially reduced by *δ*_1_ for 2 years and then by *δ*_2_:

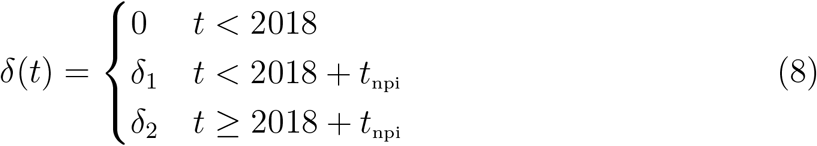

where we varied both *δ*_1_ and *δ*_2_ between 0.2 and 0.4. For each parameter set, we performed linear regression using the estimated incidence proxy as the response variable, and the predicted incidence multiplied by the scaling factor as an offset term without an intercept term—this allowed us to calculate the normal likelihood as before. For longer term predictions, we later explored further relaxing the impact of NPIs.

For the second set of assumptions, we allowed both ℛ_0_ and *ω* to vary, meaning that we no longer assumed that EV-D68 was following a biennial pattern prior to 2018. Given large geographical heterogeneity in previous EV-D68 dynamics and the underlying epidemiological parameters, we wanted to explore all possible parameter spaces and consider the possibility that EV-D68 did not follow a biennial pattern. For simplicity, we still fixed *θ* and *φ* to the same values as the first set of assumptions. In this case, we assumed that the changes in contact rates corresponds to the shape of mean Google mobility, which allowed us to simulate realistic NPI effects. Since Google mobility stopped being updated as of October 15, 2022, we assumed that the mean mobility after this period stayed constant based on the last observed value. Since we were no longer assuming a biennial epidemic, we began our simulations from 2018 and also allowed the initial proportion susceptible *S*(0) to vary across simulations; we fixed *I*(0) = 1 throughout for simplicity.

To estimate the model parameters under the second set of assumptions, we reparameterized the model to ensure the outbreak in 2018. More specifically, we let the effective reproduction number ℛ_eff_ = ℛ_0_*S*(0)/*N* to vary between 1.01 and 1.2 and the basic reproduction number to vary between 10 and 25. For simplicity, we considered the following four values of *ω* for our initial parameter search (0.1, 0.5, 1, and 2), which we found to be sufficient to obtain a good fit. For each set of parameters, we performed a linear regression using the estimated incidence proxy as the response variable against the model-predicted incidence without the intercept term.

## 3 Results

Prior to 2020, circulations of enterovirus D68 exhibited predominantly biennial patterns in the US (Fig. 1A) and in other countries. Based on this data, we previously predicted a possibility of a large outbreak in 2020, following the biennial pattern, and further showed that the decreased contact rates due to NPIs for the COVID-19 pandemic could, as observed, prevent this EV-D68 outbreak [8]. The estimated incidence proxy from the NVSN data [12] confirms our prediction: there were limited cases during 2020 followed by a large outbreak in 2022. Using the ILI time series also gave similar incidence proxy estimates (Supplementary Figure S1). Interestingly, we found signatures of continued reduction in mobility throughout 2022 (Fig. 1B), which likely affected the dynamics of EV-D68 and further contributed to delaying the timing of its resurgence.

**Figure 1:**
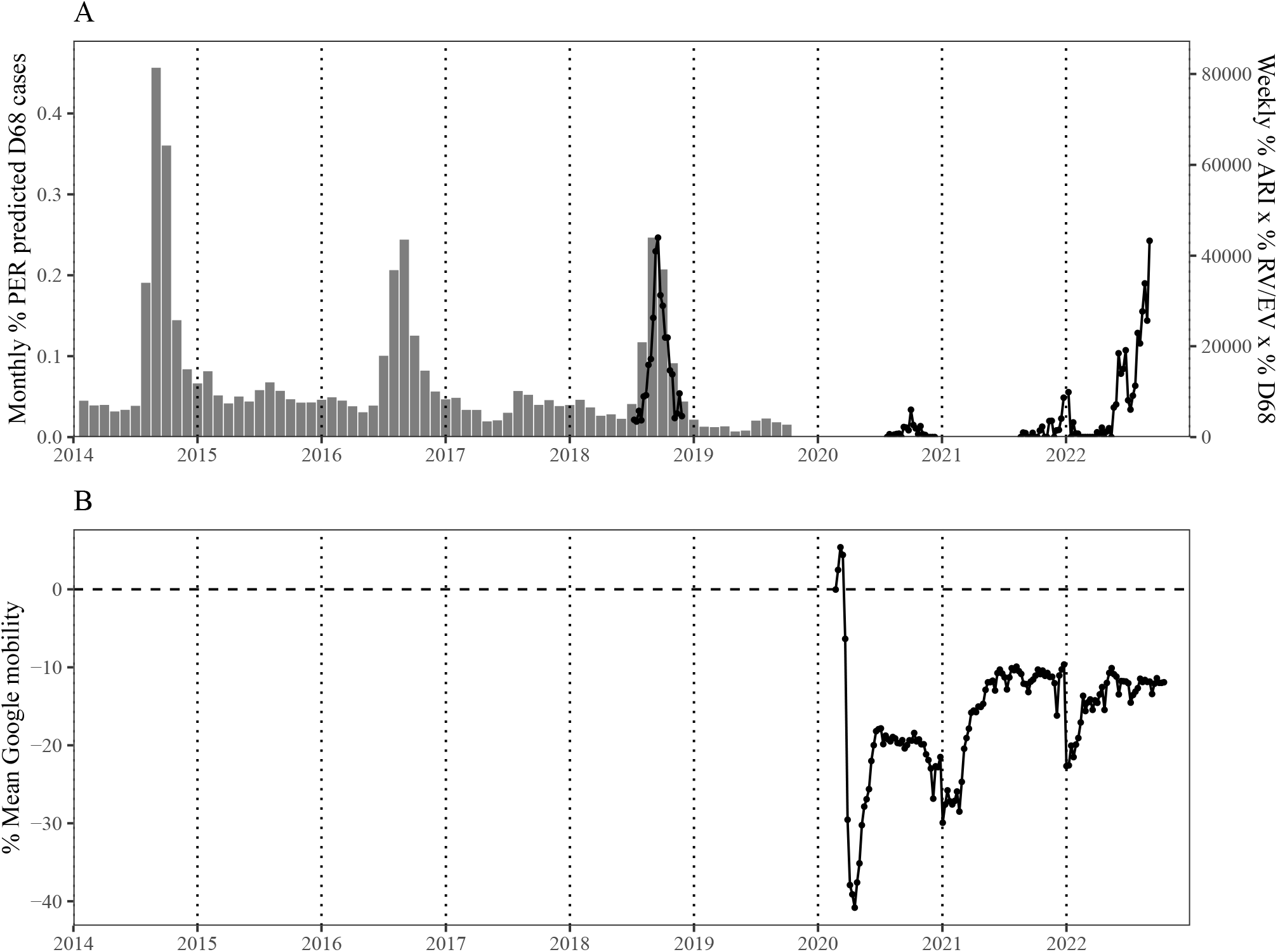
Estimated incidence proxy for enterovirus D68 circulations and Google mobility during the pandemic period in the US. (A) Weekly incidence proxy for enterovirus D68 (black lines and point) calculated by multiplying (1) weekly percentages of ED visits associated with ARIs, (2) weekly percentages of RV/EV positives, and (3) weekly percentages of EV-D68 positives. The data are digitized from the figure of [12]. Gray bars represent monthly percentages of positives for EV-D68 in the US predicted by the Pathogen Extended Resolution (PER) algorithm through the BioFire Syndromic Trends (Trend) network. The data are publicly available from [8]. (B) Percent changes in mean Google mobility across for categories (retail & recreation, grocery & pharmacy, transit stations, and workplaces). The data are publicly available from https://ourworldindata.org/covid-google-mobility-trends.

Under the first set of assumptions (endemic biennial circulation followed by step changes in contact rates), we estimated that a 28% reduction in contact rates in 2020 and 2021, followed by a 24% reduction in contact rates in 2022, provided the best fit to the observed dynamics (Fig. 2A). In this case, the model predicts a slightly larger outbreak for 2022 than for 2018. However, the likelihood surface shows a strong correlation in two parameters (i.e., the amount of reduction in transmission in 2020–2021 vs 2022; Supplementary Figure S3), meaning that many other parameter combinations can give similar fits. For example, a 40% reduction followed by a 27% reduction gives a very similar fit with *<* 2 differences in log-likelihood units. Consistent with the initial stable biennial assumption, the model predicts a return to stable biennial epidemic under a constant reduction in contact rates after 2022 (Fig. 2B); the model also predicts an increase in the overall susceptibility compared to pre-pandemic periods (Fig. 2C).

**Figure 2:**
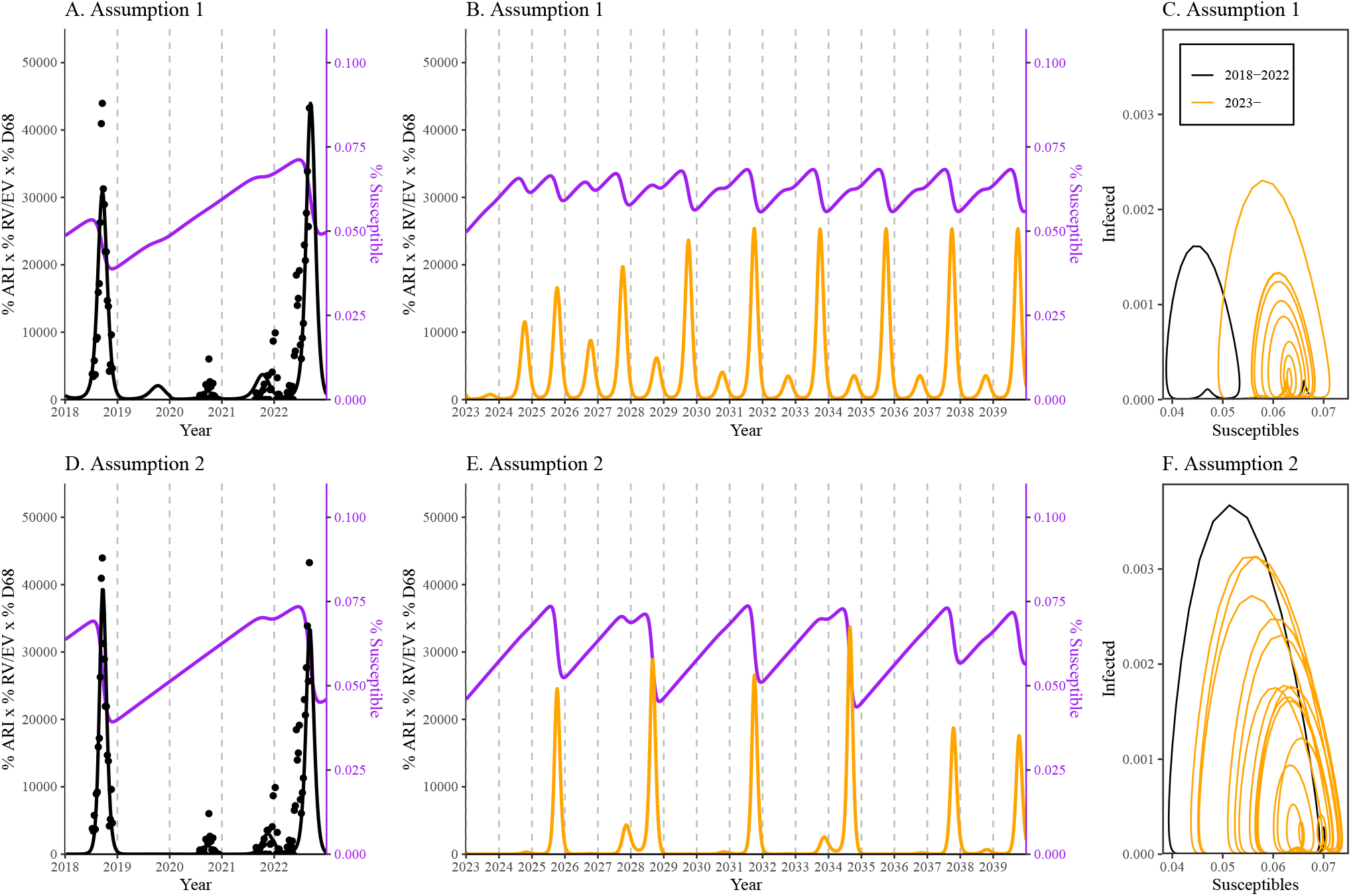
Model fits and long-term predictions under two different assumptions. (A,D) Fits of the SIR model (black lines) to incidence proxy (black points). (B,E) Longer term predictions assuming a continued reduction in contact rates. (C,F) Phase diagram of prevalence of infection (*I/N*) against the proportion susceptible (*S*/*N*).

Under the second set of assumptions (realistic changes in contact rates), we estimated ℛ_0_ = 17.5 and *ω* = 0.1 best captures the observed dynamics (Fig. 2D; Supplementary Figure S4). In this case, the model predicts a slightly larger outbreak for 2018 than for 2022. Assuming a constant reduction in contact rates after 2022, the model predicts more complex longer-term oscillatory dynamics (Fig. 2E, Fig. 2F).

Despite large uncertainties in long-term dynamics, the model predicts an absence of EV-D68 outbreak in 2023 under both sets of assumptions when there is a continued reduction in contact rates (Fig. 2A,D). However, it is likely that the contact rates will return to normal, or may have already returned to normal values. We therefore performed sensitivity analyses to characterize how short- and long-term predictions are sensitive to changes in future contact rates (Fig. 3).

**Figure 3:**
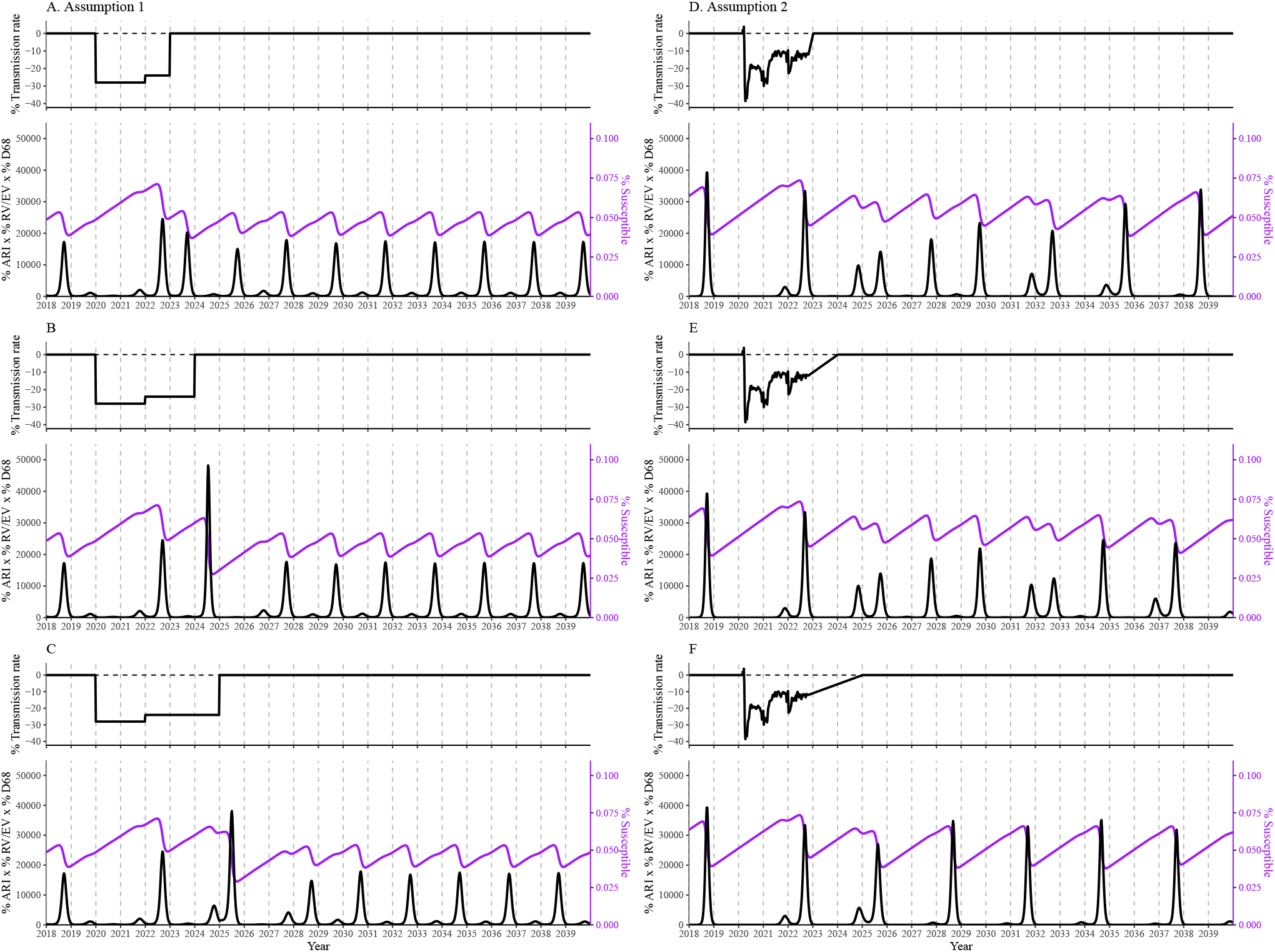
Sensitivity of long-term predictions to relaxations of changes in future contact rates. Top panels: assumed shape of the changes in contact rates *δ*(*t*) that were used to simulate the epidemics. Bottom panels: model predictions. Black lines represent the predicted incidence. Purple lines represent the susceptible dynamics. We assumed that contact rates will return to normal by the end of 2023 (A,D), 2024 (B,E), and 2025 (C,F).

Under the first set of assumptions, the model always predicts a stable biennial epidemic, but the short-term dynamics differ depending on how long it takes for contact rates to return to the pre-pandemic norm (Fig. 3A–C). Specifically, we found that an outbreak in 2023 is possible if the contact rate had returned to normal by the beginning of 2023; in all other scenarios, the model predicts no outbreak in 2023. On the other hand, we found that the impact of changes in contact rates were less predictable under the second set of assumptions (Fig. 3D–F). For example, when we assumed that the contact rates returned to normal by 2025, the predicted dynamics quickly converged to a 3 year cycle (Fig. 3F), which was not readily observed in other scenarios (Fig. 3D,E).

## 4 Discussion

In this study, we investigated the impact of COVID-19 NPIs on current and future dynamics of EV-D68 in the US. Using published data from NVSN, we reconstructed an incidence proxy for EV-D68 infections and fitted the SIR model to the incidence proxy under two different assumptions about the underlying dynamics and the shape of NPIs. For the first set of simulations, we assumed an underlying biennial epidemic and tried to estimate the shape of simple NPIs that would explain the observed dynamics. For the second set of simulations, we allowed the underlying dynamics to deviate from the biennial epidemic but instead modeled the effects of NPIs using Google mobility measures. We found that the observed dynamics can be explained under both sets of assumptions, but they predict markedly different long-term dynamics. Both sets of simulations agree in predicting the absence of large outbreak for late 2023 in most scenarios. This prediction follows intuitively from susceptible depletion following the large outbreak EV-D68 across the US. However, the potential cannot be fully ruled out: in particular, if the size of the EV-D68 outbreak in 2022 was offset by a moderate reduction in contact rates, thereby preventing a significant susceptible depletion, a sudden increase in contact between 2022 and 2023 can still cause an outbreak in 2023. Serological surveys would help reduce uncertainties in both data and model predictions [17].

The two sets of assumptions that we considered represent different extremes. The first captures the biennial patterns of EV-D68 outbreaks but makes simplifying assumptions about the impact of NPIs. The second set of assumptions captures realistic changes in contact rates but does not account for the dynamics before 2018. Given large uncertainties in EV-D68 dynamics before 2018 and the exact changes in contact rates after the emergence of COVID-19, it is difficult to assess which assumptions are more realistic. We tried combining a biennial pattern with realistic NPIs but such a combination did a poor job at capturing the observed dynamics. Therefore, our analysis highlights uncertainties in both short- and long-term dynamics of EV-D68. Overall, more detailed surveillance is required to better understand the dynamics of EV-D68.

There are many limitations to our analysis. First of all, surveillance efforts for EV-D68 are currently limited in the US, adding large uncertainties to its observed dynamics. Even though EV-D68 is thought to have circulated every two years before 2020, earlier analysis revealed large geographical heterogeneity in its patterns of spread. Therefore, our estimated incidence proxy for the US provides a crude view of the overall dynamics and does not capture any geographical heterogeneity that may be present. Second, we did not have access to the raw data and so we manually digitized the data. This procedure likely added inaccuracies to the data (though this is likely not a major error). Finally, our mathematical model necessarily relies on many simplifying assumptions that neglect age heterogeneities, immune and phylodynamic complexities, etc. However, it would be impractical to try to fit more complicated models at this juncture, due to limited data.

There are considerable uncertainties in model parameters that we did not consider in our analysis. Instead, we assumed that many parameters were known prior to model fits. Nonetheless, our analysis already demonstrates that a wide range of scenarios are consistent with the observed dynamics, and these scenarios give vastly different predictions. Further exploring parameter spaces would provide a broader overview of potential epidemic trajectories, but our qualitative conclusion about the uncertainties around future dynamics would not change.

Given the uncoupling of EV-D68 circulation with AFM cases in 2022, uncertainty remains about the frequency of AFM cases associated with future outbreaks of particular strains of EV-D68. It is unclear at this time whether changes in the virus, changes in host immunity, or both caused the apparent lack of AFM in 2022 despite the large EV-D68 outbreak. For example, EV-D68 strains that circulated in 2022 are broadly similar to those in 2018, but even small changes in the virus can lead to large changes in neurovirulence. On the other hand, an increase in the susceptible pool during the pandemic could have also caused an increase in the mean age of infection of EV-D68 in 2022, which may lead to less severe infections. A coupling of molecular epidemiology and detailed surveillance data, such as age-structured surveillance or serological surveys, may provide better power to test different hypotheses and understand the relationship between AFM and EV-D68. Further extending the analysis to EV-D68 outbreak dynamics in other countries and massive natural experiments of parallel impacts of COVID-19 NPI measures to dynamics of other endemic viral infections is critical [9].

## Data Availability

All data used in this study are publicly available.

## Acknowledgements

We thank the NIH, VRC, PREMISE Program, Claire Midgley, Janell A. Routh, and Heidi Moline for help and discussions. We thank the New Vaccine Surveillance Network (NVSN) for generating, and providing summaries of, key EV-D68 surveillance data.

## Supplementary Materials

**Figure S1:**
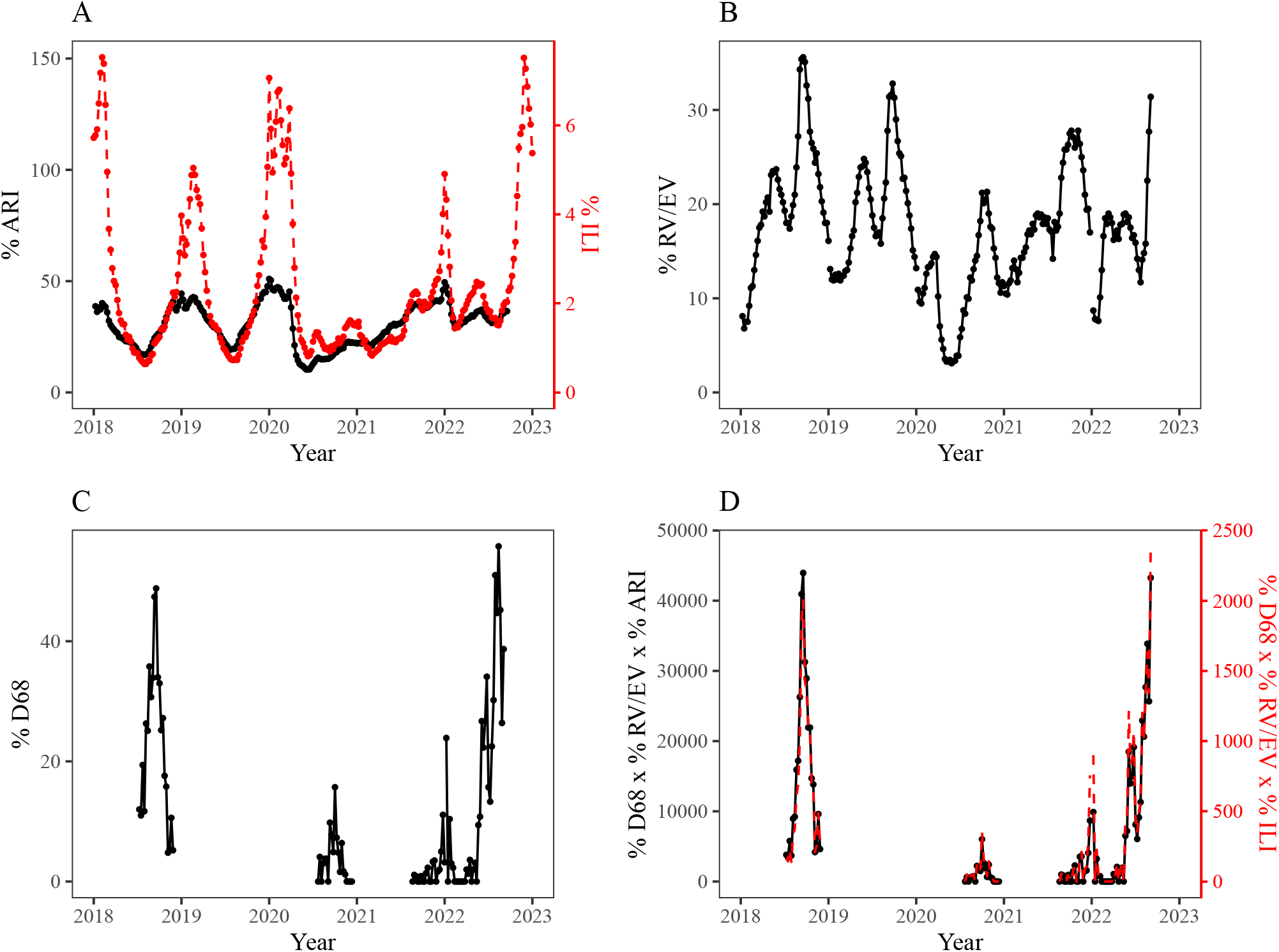
Time series representing circulations of respiratory diseases and EV-D68. (A) Weekly percentages of ED visits for children and adolescents (¡18 years old) associated with ARIs (black) and the weekly proportions of ILI visits (red). (B) Weekly percentages of positives for rhinovirus and enterovirus. (C) Weekly percentages of positives for EV-D68 among RV/EV positives. (D) Weekly incidence proxy estimated using ARIs (black) and ILI (red) time series.

**Figure S2:**
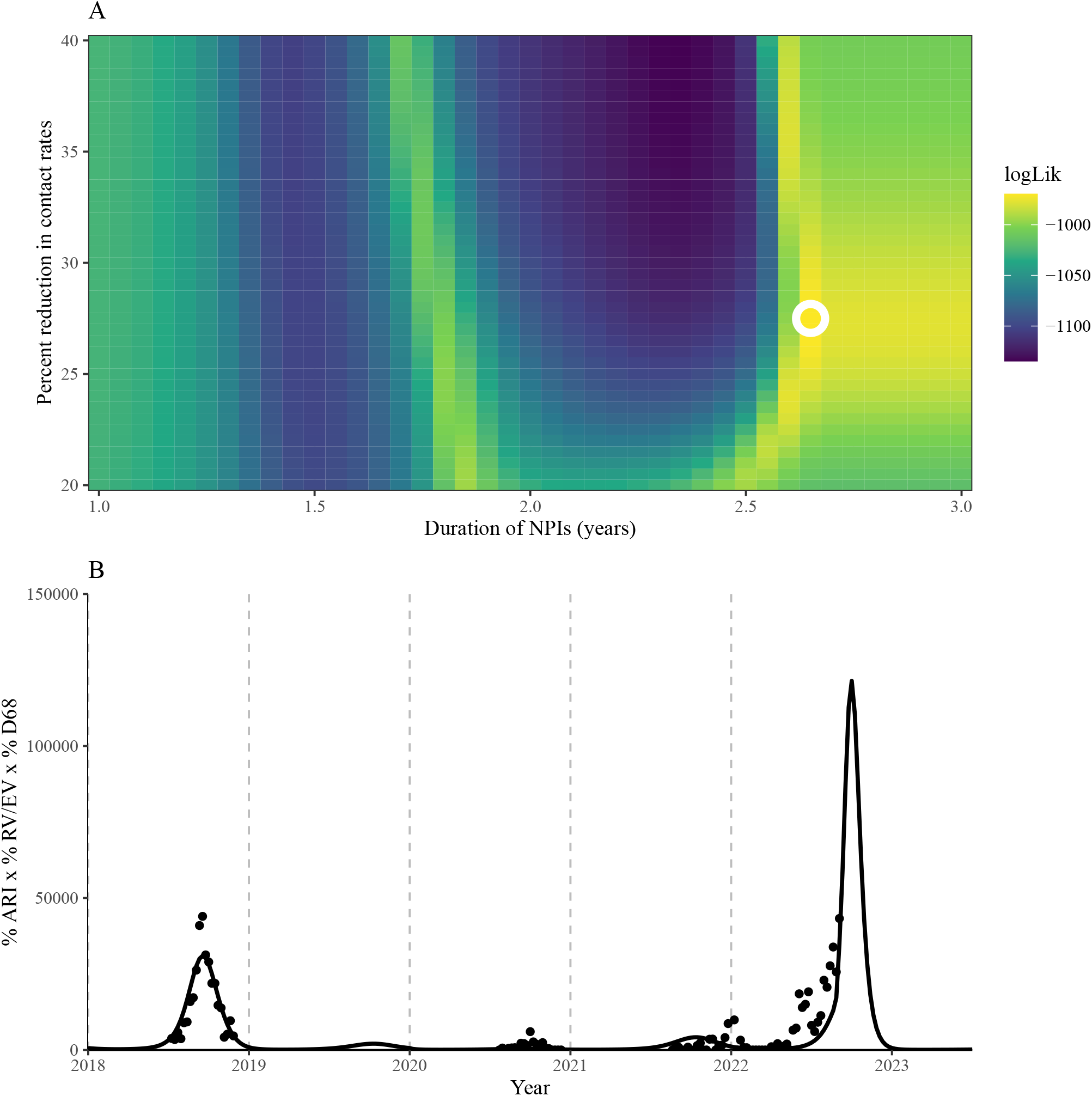
Results of fitting the SIR model with a constant reduction in contact rates over a fixed period. (A) The log likelihood surface of the model fits. The circle represents the best fitting parameter. (B) The best fit of the SIR model (black lines) to incidence proxy (black points).

**Figure S3:**
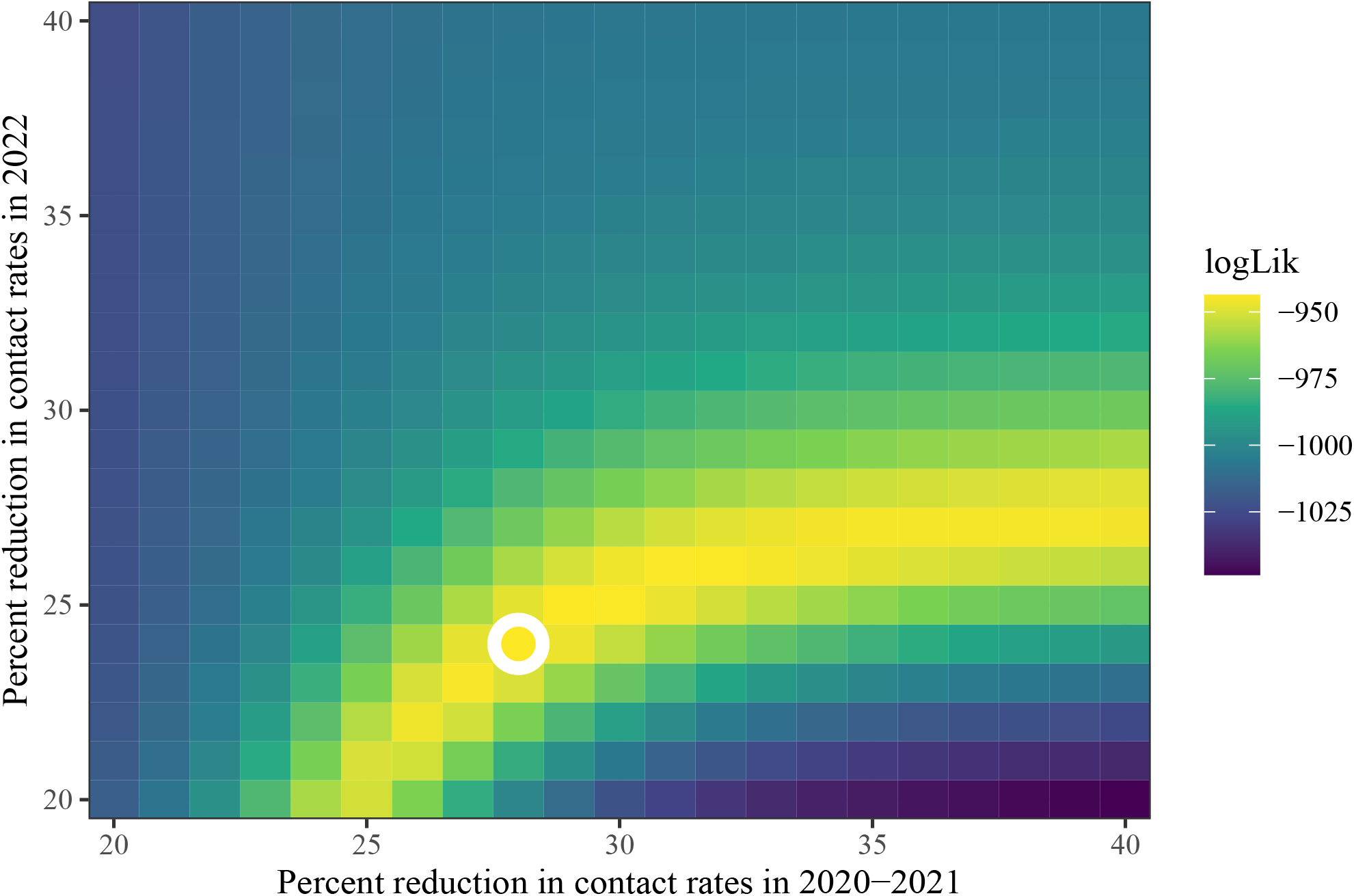
The log likelihood surface of the model fits under the first set of assumptions. The circle represents the best fitting parameter.

**Figure S4:**
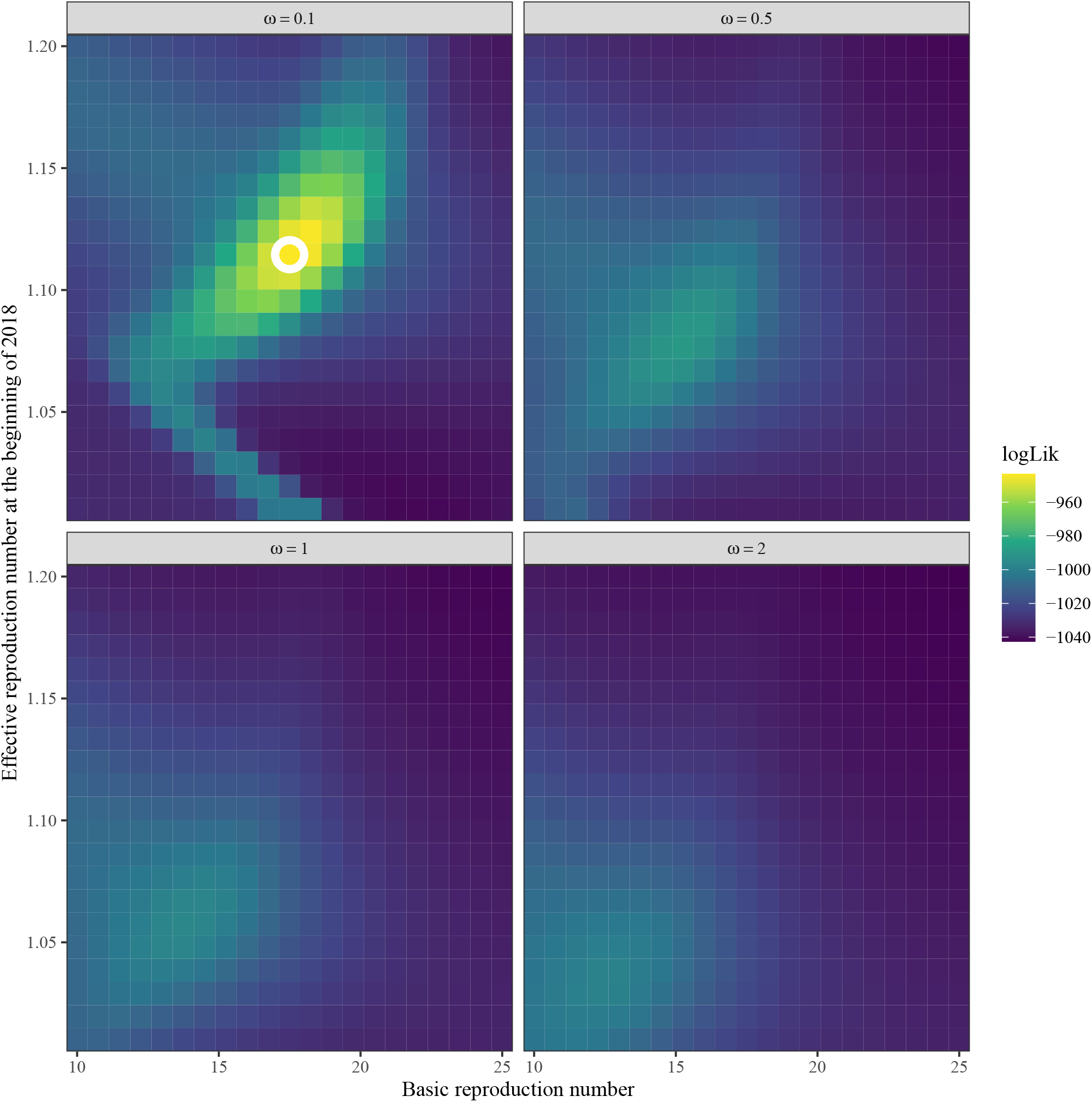
The log likelihood surface of the model fits under the second set of assumptions. The circle represents the best fitting parameter.

